# Assessment of Right Ventricular Endocardial Fibroelastosis in Fetuses with Critical Pulmonary Stenosis and Pulmonary Atresia with Intact Ventricular Septum

**DOI:** 10.1101/2023.08.04.23293682

**Authors:** Yue Wang, Gang Luo, Yi Sun, Taotao Chen, Silin Pan

## Abstract

**Background:** Systematic assessment of right ventricular (RV) endocardial fibroelastosis (EFE) in the fetus with critical pulmonary stenosis and pulmonary atresia with intact ventricular septum (CPS/PA-IVS) has not been reported. The implications of RV EFE for circulatory outcomes with or without fetal cardiac intervention (FCI) have also not been demonstrated.

**Methods:** The fetal echocardiographic data from July 2018 to January 2021 in a single-center institution were collected. Three reviewers independently graded EFE according to the presence and extent of endocardial echogenicity. The associations among EFE severity, anatomic variables, and postnatal outcomes were analyzed.

**Results:** 81 cases with RV EFE were identified. By consensus, EFE severity was assessed as grade 1 (“mild”; n=66, 81.48%), grade 2 (“moderate”; n=11, 13.58%), and grade 3 (“severe”; n=4, 4.94%). RV sphericity values were greater in grade 2 and 3 EFE groups compared to the grade 1 EFE group, implying more severe noncompliance and worse diastolic function. 10/81(12.35%) fetuses underwent FCI. At the latest available follow-up, 59/81 (72.84%) patients had achieved biventricular circulation. Furthermore, 9/59 (15.25%)—four with grade 1 EFE, three with grade 2, and two with grade 3—had undergone FCI, whereas the other 50/59 (84.75%)—who all had grade 1 EFE—had not.

**Conclusions:** For the first time in CPS/PA-IVS, RV EFE was graded and the potential of FCI to improve the prognosis of non-mild RV EFE patients was revealed. Non-mild RV EFE was suggested to be a putative indicator of FCI.

**Graphic Abstract:** 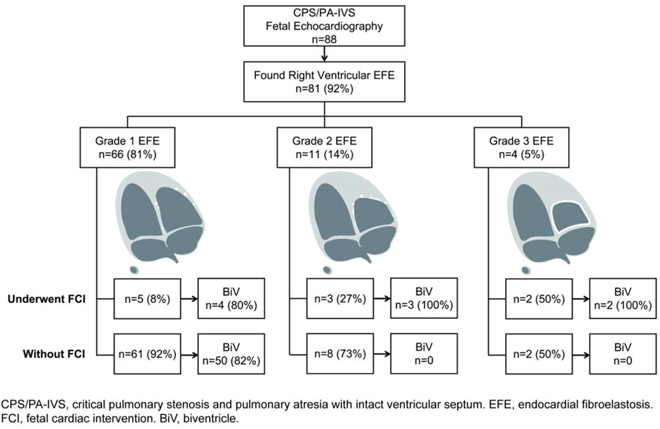

## What is Known

In critical pulmonary stenosis and pulmonary atresia with intact ventricular septum (CPS/PA-IVS), an appropriate fetal cardiac intervention (FCI) can increase the probability of a biventricular outcome.

The FCI indications in CPS/PA-IVS remain debatable.

FCI indications mainly include geometric and morphological indicators, while pathophysiologic variables are rarely assessed before FCI.

## What the Study Adds

For the first time in CPS/PA-IVS, right ventricular endocardial fibroelastosis is systematically assessed as mild, moderate, or severe according to the endocardial echogenicity.

This single-center cohort study demonstrates the potential of FCI to improve the prognosis of individuals with non-mild endocardial fibroelastosis.

Right ventricular non-mild endocardial fibroelastosis is suggested to be a putative indicator of FCI.

## Introduction

Pulmonary atresia with intact ventricular septum (PA-IVS), characterized by right ventricular antegrade blood flow obstruction, is a rare form of complex congenital heart disease. Critical pulmonary stenosis (CPS) resembles the favorable extreme circumstance of PA-IVS. Both CPS and PA-IVS are severe right ventricular outflow tract (RVOT) obstructions, that have analogous hemodynamic alterations.^1^ Associated neonates require repeated postnatal catheter intervention and surgery and struggle to achieve biventricular circulation^2^, resulting in an exceedingly poor prognosis.

To optimize the conditions for right ventricle (RV) and valves growth in utero, the RVOT obstruction need to be eliminated. A successful fetal cardiac intervention (FCI) can relieve the RVOT obstruction and promote continued growth of RV during the prenatal period, increasing the probability of a biventricular outcome. CPS/PA-IVS patients have been reported to have improved postnatal prognoses following FCI.^3, 4^ Successful FCIs have been done in our hospital in the last five years, and the outcomes are exceptional.^5–7^

However, FCI indications in CPS/PA-IVS remain debatable. Case selection is critical and challenging due to the limited population and scarcity of data. Most institutions base their decision on geometric and morphological indicators, such as RV structures in respect to the left sided structures, and indications of RV preload and pressure-generating capacity.^4, 8^ Despite meeting the current FCI criteria, some patients achieved biventricular outcomes without FCI.^5, 9^ Due to the limited population of CPS/PA-IVS, restricted examination technology, ethical considerations, and other factors, the pathophysiologic variables are rarely assessed before FCI. More comprehensive and systematic assessing criteria need to be developed.

Endocardial fibroelastosis (EFE), which is represented by an abnormal collagen and elastic fiber distribution in the endocardial layer of ventricles, has been proven to hinder pre- and postnatal heart development. Under pathological conditions, EFE can be detected in all four heart chambers, but primarily in the left ventricle. Due to its inelastic fibrotic attributes, EFE in ventricles is hypothesized to produce relative diastolic noncompliance and restrict heart growth. Compliance deteriorates when the RV becomes more fibrotic and hypertrophic, which worsens ventricular diastolic dysfunction. Therefore, development of heart both before and after birth may be hindered by EFE. The RV myocardium, on the contrary, could additionally sustain secondary damage as a consequence of CPS/PA-IVS. Additional hemodynamic disturbances would result from a hypoplastic right ventricle.^10^ During heart development, abnormal hemodynamic loading will also stimulate the production of collagen and elastin in the endocardium.

In CPS/PA-IVS, tricuspid valve and pulmonary valve deficits lead to flow disturbances manifested as jets across the valve onto localized regions of the RV cavity, which in most cases increases the pressure in the RV. In response to the elevated RV pressure, EFE develops around the endocardium and thickens it. In hypoplastic left heart syndrome (HLHS) diseases, EFE has been established to play a vital role in disease progression^10^. The incorporation of EFE severity into a threshold scoring system evaluating potential for biventricular circulation for midgestation fetuses with aortic stenosis and evolving HLHS enhanced the sensitivity and positive predictive value for postnatal outcomes.^11^ However, due to the notable heterogeneity and smaller population, no published data are available regarding the reliability of systematic grading of RV EFE in CPS/PA-IVS fetuses.

It was assumed, based on clinical practice, that RV EFE might substantially influence circulatory outcomes of CPA/PA-IVS and, therefore, be a novel FCI indicator. The current study aimed to grade RV EFE severity in CPS/PA-IVS and discover the relationship between itself and morphological parameters, assisting in finding more appropriate and comprehensive FCI indications.

## Methods

### Study Cohort

The retrospective single-center cohort study included fetuses with CPS/PA-IVS from July 2018 to January 2021 in Women and Children’s Hospital, Qingdao University. Patients who had ceased gestation or lacked follow-up records were eliminated.

This study was approved by Ethics Committee of Medical College of Qingdao University (QDU-HEC-2022142). Each participant’s legal guardian provided written informed consent prior to enrolling in the study. All studies were conducted in accordance with both the Declarations of Helsinki and Istanbul.

### Analysis of Echocardiographic Images

The International Society of Ultrasound in Obstetrics and Gynecology guidelines were followed for all fetal echocardiography.^12^ Echocardiography was performed with Siemens Acuson Sequoia (Siemens Medical Solutions USA, Inc., Pennsylvania) and Philips Compact 5000 (Philips Healthcare, Massachusetts) equipment, and the setting parameters were modified to optimize image quality in accordance with our hospital’s standard procedure. Two fetal echocardiographers and an experienced cardiologist who were unaware of the postnatal outcome or whether the fetuses had undergone FCI reviewed the echocardiographic pictures and videos. Each study variable was assessed by consensus. Four-chamber view, RV outflow view, left ventricular (LV) outflow view, three vessels and trachea view, and other views were analyzed.

The ventricular and valvular dimensions were measured, including RV long-axis dimension, RV short-axis dimension, LV long-axis dimension, pulmonary valve (PV) annulus diameter, aortic valve (AV) annulus diameter, tricuspid valve (TV) annulus diameter, and mitral valve (MV) annulus diameter. Right ventricular sphericity was estimated as the ratio of short-axis to long-axis dimension.

Based on clinical experience, it was assumed that RV EFE could substantially impact circulatory outcomes and, therefore, be a potential FCI indicator. Due to the absence of an established grading standard in RVOT obstruction diseases, we strived to systematically assess the RV EFE. The EFE was repeatedly assessed mainly on the basis of echocardiographic findings of the presence, extent, and range of endocardial brightness (echogenicity) in RV. In addition, the EFE within the papillary muscles and the septal EFE were included.^13, 14^

RV EFE gradation in the present study is based primarily on the experience of echocardiography reviewers. RV EFE severity in CPS/PA-IVS was graded as: 1, mild (scattered echogenic spots within the RV, including the TV and papillary muscles); 2, moderate (noncontiguous echogenic patches throughout the RV); and 3, severe (contiguous echogenic lining of the RV). **Figure 1** shows the examples of each RV EFE grade.

**Figure 1.**
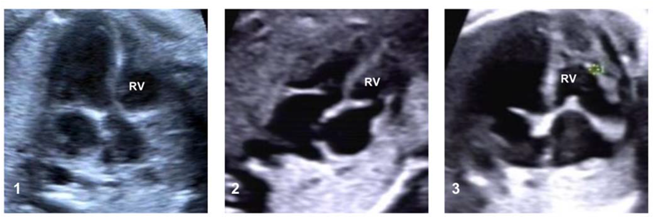
Echocardiographic Images Demonstrating Examples of RV EFE Grade 1, 2, and 3. RV, right ventricle; EFE, endocardial fibroelastosis.

### FCI Procedure

In CPS/PA-IVS, the method of FCI is fetal pulmonary valvuloplasty (FPV) under ultrasound. FPV procedures were previously described in detail.^5^ In our hospital, FCI was considered in following situations: membranous PA, an intact or highly restrictive ventricular septum, and hypoplastic valves with a visible but relatively small RV. The exclusion criteria include muscular atresia of the RVOT, severe fetal edema, severe tricuspid regurgitation with low velocity (<2.5 m/s), and right ventricle-dependent coronary circulation.

### Data Collection

Data of maternal and fetal features, procedure records, and postnatal outcomes of fetuses diagnosed with CPS/PA-IVS were collected. No sex-based or race/ethnicity-based differences were included. The corresponding author had full access to all the data in this study and takes responsibility for its integrity and the data analysis.

Postnatal outcomes were classified as biventricular (BiV) circulation and non-BiV circulation. According to the established standards, BiV was defined as the RV is the only source of pulmonary blood flow.^3, 15^ It included individuals without an atrial septal defect or with an atrial septal defect shunting predominantly left-to-right and saturations of 92% at the latest follow-up.^16^ Non-BiV circulatory patterns included univentricular and one-and-a-half circulation.

### Statistics

One-way ANOVA was used to analyze the relationship between the EFE gradation and other variables. Tamhane multiple comparison approach was used to compare the components in different EFE grades. A p-value <0.05 is considered statistically significant.

## Results

### Demographical Characteristics

Between July 2018 to January 2021, a total of 178 consecutive cases were collected from our institution. After excluding patients who ceased gestation (n=78) or lacked follow-up records (n=12), 88 echocardiographic recordings were reviewed. 81/88 (92.05%) with RV EFE were found.

10/81 fetuses underwent FPV and all procedures were performed successfully without severe complications. And the echocardiograms before FCI were used to compare with those of non-FCI fetuses. FCI were considered technically successful if a balloon was inflated across the valve and there was improvement in anterograde flow without severe complications, as defined previously.^3^

### Echocardiographic Findings

By consensus, the EFE severity was assessed as grade 1 in 66 (81.48%), grade 2 in 11 (13.58%), and grade 3 in 4 (4.94%). In the 10 fetuses underwent FPV, five were assessed as grade 1 EFE, three as grade 2 EFE and two as grade 3 EFE. In the 71 fetuses who didn’t undergo FPV, 61 (85.92%) were assessed as grade 1 EFE, 8 (11.27%) as grade 2 EFE, and 2 (2.81%) as grade 3 EFE.

Data in **Table 1** and **Figure 2** showed a statistically significant difference in RV sphericity among the three groups (p<0.001). The grade 2 and 3 EFE groups had larger RV sphericity values (RV short-axis to long-axis dimension) than the grade 1 EFE group, with statistically significant differences (p<0.001 and p=0.04, respectively). Between grade 2 and 3 EFE groups, there is no discernible difference in RV sphericity (p=0.6). Notably, larger RV sphericity implied more severe noncompliance and worse diastolic function.

**Table 1.**
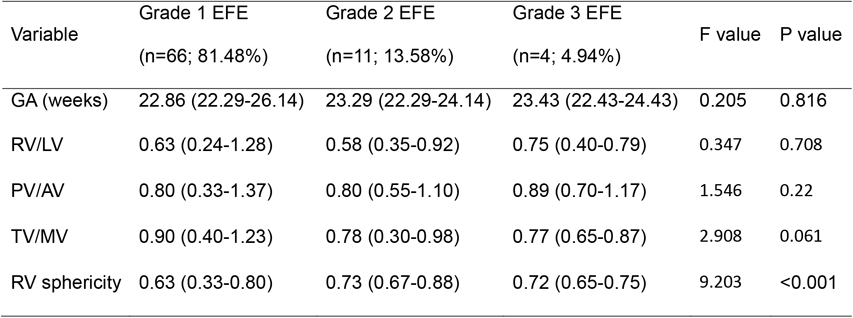
Anatomic Characteristics on Fetal Echocardiography.

**Figure 2.**
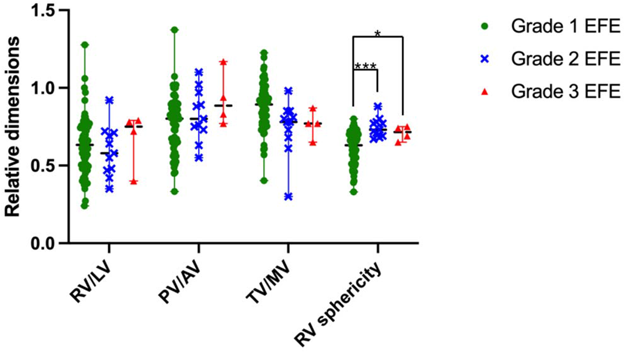
Anatomical Characteristics of Individual Fetuses. Anatomical characteristics from 81 fetal echocardiography were presented. RV sphericity was estimated as the right ventricular short-axis to long-axis dimension ratio. EFE, endocardial fibroelastosis; RV/LV, right/left ventricular long-axis dimension; PV/AV, pulmonary/aortic valve annulus diameter; TV/MV, tricuspid/mitral valve annulus diameter; *, p<0.05; ***, p<0.001.

Gestational age, RV/LV, PV/AV, and TV/MV were not statistically different among the three groups (p>0.05).

### Procedures and Postnatal Circulatory Outcomes

The average follow-up time was 32 months. Till the end of follow-up, 59/81 (72.84%) patients have achieved BiV circulation. And 9/59 (15.25%)—four with grade 1 EFE, three with grade 2 EFE, and two with grade 3 EFE—had undergone FCI, whereas the other 50/59 (84.75%)—who all had grade 1 EFE—had not (**Table 2**). 22/81 (27.16%) patients—twelve with grade 1 EFE, eight with grade 2 EFE, and two with grade 1 EFE—haven’t achieved BiV circulation (**Table 2**). One case with grade 1 EFE had undergone FCI but not achieved a BiV outcome, probably due to relatively severe hypoplastic RV and valves (RV/LV=0.47, PV/AV=0.65, TV/MV=0.76, RV sphericity=0.43).

**Table 2.** Pre- and Postnatal Procedures and Circulatory Outcomes.

| Outcome | Procedure | Number of cases |  |  |
| --- | --- | --- | --- | --- |
|  |  | Grade 1 EFE | Grade 2 EFE | Grade 3 EFE |
| BiV (n=59; 72.84%) |  |  |  |  |
|  | FCI + postnatal interventions<br>(n=9; 15.25%) | 4 | 3 | 2 |
|  | Non-FCI + postnatal interventions<br>(n=50; 84.75%) | 50 | 0 | 0 |
| Non-BiV (n=22; 27.16%) |  |  |  |  |
|  | FCI + postnatal interventions<br>(n=1; 4.55%) | 1 | 0 | 0 |
|  | Non-FCI + postnatal interventions with<br>or without univentricular palliation<br>(n=21; 95.45%) | 11 | 8 | 2 |

The results indicated that FCI had certain potential to improve postnatal circulatory outcomes in fetuses with non-mild (grade 2 and 3) EFE. Non-mild RV EFE was suggested to be a putative indicator of FCI.

## Discussion

In the present study, three grades of RV EFE in CPS/PA-IVS fetuses were defined for the first time. To the best of our knowledge, this is the first report about RV EFE severity grading in RVOT obstruction diseases. In the current study, the results revealed that RV sphericity values were greater in grade 2 and 3 EFE groups compared to the grade 1 EFE group, indicating the association of EFE severity with RV sphericity. As the normal RV is elliptical, stiffness and hypoplasia of the RV might manifest as increased sphericity.^17^ Meanwhile, there was no statistically significant difference among the three groups in terms of RV/LV, PV/AV, or TV/MV (p>0.05). These results suggested that the severity of EFE was an independent risk factor that had no significant impact on the anatomical parameters mentioned above, consistent with HLHS.^11^

RV EFE severity was also associated with the postnatal outcome. Without a successful FCI, individuals with grade 2 and 3 EFE were more likely to achieve non-BiV circulations. Fetuses with grade 1 EFE, in contrast, had a more favorable prognosis and were more likely to achieve biventricular outcomes without FCI.

The presence and severity of EFE have been suggested as including criteria for intrauterine balloon valvuloplasty in fetuses with critical aortic stenosis, and they relate with the possibility of a biventricular outcome following prenatal intervention for fetal aortic stenosis with evolving HLHS.^11, 13^ In up to 70% HLHS, EFE limits the left ventricular outflow tract (LVOT) .^18^ However, systematic assessment of RV EFE has not been reported.

The goal of FCI is to enhance the possibility of a biventricular circulation so as to improve long-term outcomes. Ventricular and valvular hypoplasia, fibrosis, and abnormalities in coronary artery are prominent predictors of poor outcomes in individuals with CPS/PS-IVS. The current study demonstrated successful FCI could potentially be capable to halt the progression of RV hypoplasia in fetuses with non-mild (grade 2 or 3) EFE. It has been proven that in the early stages of LVOT disorders, the fibrotic alterations are reversible.^19^ Since fibrosis in the setting of RV hypertension is known to be partially reversible in postnatal congenital heart disease (such as tetralogy of Fallot), cautiously indicated and timed prenatal intervention may be able to prevent the development of excessive EFE, which could otherwise seriously impair RV function and result in single ventricular circulation and a poor prognosis.^19^ FCI should be taken into account if there is any potential to restore RV development and preserve a biventricular outcome. Furthermore, FCI has demonstrated benefits in improving prognosis. Nevertheless, the current FCI for CPS/PA-IVS criteria were still inconsistent and contradictory among institutions.^8^

Our institution has done successful FCIs in the last five years.^5, 6, 20^ And our team is well-trained in perinatal medicine, including ultrasound diagnosis, maternal anesthesia, catheter intervention, complication management, and follow-up investigation. It has come into focus that despite matching the current FCI standards, some patients acquired biventricular circulation without FCI.^5, 9^ Thus, for the purpose of more accurately assessing FCI indication, we intended to identify a new predictor. Fortunately, non-mild RV EFE was found to be a potent indicator for FCI.

Clarifying the theoretical mechanisms and natural progression of CPS/PA-IVS is crucial to guide appropriate patient selection and timing for FCI. In the developing heart, EFE secondary to altered hemodynamics has been discovered.^19^ And when EFE occurs, it is directly related to flow disruptions and presents an infiltrative growth pattern, increasing the diastolic stiffness of RV. Aging promotes structural changes in EFE that lead to decreased cellularity and growth that is more infiltrative into the myocardium beneath.^10^ Endothelial-to-mesenchymal transition (EndMT) has been demonstrated as one underlying mechanism of EFE formation. Endothelial cells of the endocardial layer convert into fibroblasts as a consequence of EndMT.^21^ Patients with stenotic or defective valves that cause flow disturbances in the ventricles developed fibrotic tissue covering the ventricle resulting from EndMT. Thus, EndMT may be a potential therapeutic traget.

There exists several limitations in the present study. First and foremost, the integrative effect of EFE together with other predictors on postnatal outcomes remains unclear. Also, the follow-up duration was relatively short. Although the short-term data are promising, further study is necessary for long-term data. Additionally, the accuracy of the echocardiographic assessment of EFE are not flawless.^22, 23^ The “EFE” assessed in the present study was actually echocardiographic echodensity, which was assumed to result from EFE. This is an assumption that no standard could verify because fetal autopsy was available in only 2 of 178 cases. These two fetuses both exhibited grade 2 EFE on echocardiography and fibrosis was confirmed by pathologic findings in their autopsy samples. What’s more, the acquisition of a proper fetal echocardiographic view is challenging due to plenty of factors, such as the tiny size of the fetal heart, rapid heart rate, and frequent fetal motion.^22^ Despite these limitations, the present work has great significance for systematically assessing RV EFE in CPS/PA-IVS. Non-mild RV EFE was suggested to be a putative indicator of FCI.

In summary, we graded RV EFE in CPS/PA-IVS and revealed the potential of FCI to improve the prognosis of non-mild EFE patients for the first time. Further study is warranted to integrate EFE with other predictors in order to more appropriately select patients.

## Data Availability

All data referred to in the manuscript are available from the corresponding author upon reasonable request.

## Acknowledgements

None.

## Non-standard Abbreviations and Acronyms

AV: aortic valve
BiV: biventricle
CPS: critical pulmonary stenosis
EFE: endocardial fibroelastosis
EndMT: endothelial-to-mesenchymal transition
FCI: fetal cardiac intervention
FPV: fetal pulmonary valvuloplasty
HLHS: hypoplastic left heart syndrome
LV: left ventricle
LVOT: left ventricular outflow tract
MV: mitral valve
PA-IVS: pulmonary atresia with intact ventricular septum
PBPV: percutaneous balloon pulmonary valvuloplasty
PDA: patent ductus arterios
PV: pulmonary valve
RV: right ventricle
RVOT: right ventricular outflow tract
TV: tricuspid valve.

## References

1. Kwiatkowski DM, Hanley FL, Krawczeski CD. Right ventricular outflow tract obstruction: pulmonary atresia with intact ventricular septum, pulmonary stenosis, and Ebstein’s malformation. Pediatr Crit Care Med. 2016;17:S323–329. doi: 10.1097/PCC.0000000000000818

2. Shahanavaz S, Qureshi AM, Petit CJ, Goldstein BH, Glatz AC, Bauser-Heaton HD, McCracken CE, Kelleman MS, Law MA, Nicholson GT, et al. Factors influencing reintervention following ductal artery stent implantation for ductal-dependent pulmonary blood flow: results from the congenital cardiac research collaborative. Circ Cardiovasc Interv. 2021;14:e010086–010095. doi: 10.1161/CIRCINTERVENTIONS.120.010086

3. Tworetzky W, McElhinney DB, Marx GR, Benson CB, Brusseau R, Morash D, Wilkins-Haug LE, Lock JE, Marshall AC. In utero valvuloplasty for pulmonary atresia with hypoplastic right ventricle: techniques and outcomes. Pediatrics. 2009;124:e510–518. doi: 10.1542/peds.2008-2014

4. Hogan WJ, Grinenco S, Armstrong A, Devlieger R, Dangel J, Ferrer Q, Frommelt M, Galindo A, Gardiner H, Gelehrter S, et al. Fetal cardiac intervention for pulmonary atresia with intact ventricular septum: international fetal cardiac intervention registry. Fetal Diagn Ther. 2020:1–9. doi: 10.1159/000508045

5. Luo G, Gao S, Sun HX, Ji ZX, Wang DL, Sun Y, Pan SL. Valvuloplasty of fetal pulmonary atresia with intact ventricular septum and hypoplastic right heart: Mid-term follow-up results. J Interv Med. 2022;5:196–199. doi: 10.1016/j.jimed.2022.04.001

6. Xing QS, Sun Y, Luo G, Zhang A, Chen TT, Pan SL. Intrauterine intervention of pulmonary atresia at 26(th) gestational week. Chin Med J (Engl). 2018;131:2880–2881. doi: 10.4103/0366-6999.246074

7. Luo G, Liu N, Wang KL, Sun Y, Zhang A, Pan SL. Immediate and short-time outcomes of pulmonary valvuloplasty in a fetus with pulmonary atresia. Chin Med J (Engl). 2019;132:1758–1759. doi: 10.1097/CM9.0000000000000322

8. Villalain C, Moon-Grady AJ, Herberg U, Strainic J, Cohen JL, Shah A, Levi DS, Gomez-Montes E, Herraiz I, Galindo A. Prediction of postnatal circulation in pulmonary atresia/critical stenosis with intact ventricular septum: systematic review and external validation of models. Ultrasound Obstet Gynecol. 2023;62:14–22. doi: 10.1002/uog.26176

9. Sun HX, Luo G, Wang SB, Ji ZX, Chen TT, Pan SL. [Postnatal management and follow-up of six fetuses affected by pulmonary atresia with intact ventricular septum and right ventricular hypoplasia without intrauterine intervention]. Chin J Perinat. 2022;25:576–581. doi:10.3760/cma.j.cn113903-20211111-00945

10. Weixler V, Marx GR, Hammer PE, Emani SM, Del Nido PJ, Friehs I. Flow disturbances and the development of endocardial fibroelastosis. J Thorac Cardiovasc Surg. 2020;159:637–646. doi: 10.1016/j.jtcvs.2019.08.101

11. McElhinney DB, Vogel M, Benson CB, Marshall AC, Wilkins-Haug LE, Silva V, Tworetzky W. Assessment of left ventricular endocardial fibroelastosis in fetuses with aortic stenosis and evolving hypoplastic left heart syndrome. Am J Cardiol. 2010;106:1792–1797. doi: 10.1016/j.amjcard.2010.08.022

12. International Society of Ultrasound in Obstetrics and Gynecology, Carvalho JS, Allan LD, Chaoui R, Copel JA, DeVore GR, Hecher K, Lee W, Munoz H, et al. ISUOG practice guidelines (updated): sonographic screening examination of the fetal heart. Ultrasound Obstet Gynecol. 2013;41:348–359. doi: 10.1002/uog.12403

13. Axt-Fliedner R, Tenzer A, Kawecki A, Degenhardt J, Schranz D, Valeske K, Vogel M, Kohl T, Enzensberger C. Prenatal assessment of ventriculocoronary connections and ventricular endocardial fibroelastosis in hypoplastic left heart. Ultraschall Med. 2014;35:357–363. doi: 10.1055/s-0034-1366361

14. Jiang Y, Xu Y, Tang J, Xia H. Assessment of structural and functional abnormalities of the myocardium and the ascending aorta in fetus with hypoplastic left heart syndrome. Biomed Res Int. 2016;2016:2616729–2616737. doi: 10.1155/2016/2616729

15. Donofrio MT, Moon-Grady AJ, Hornberger LK, Copel JA, Sklansky MS, Abuhamad A, Cuneo BF, Huhta JC, Jonas RA, Krishnan A, et al. Diagnosis and treatment of fetal cardiac disease: a scientific statement from the American Heart Association. Circulation. 2014;129:2183–2242. doi: 10.1161/01.cir.0000437597.44550.5d

16. Petit CJ, Glatz AC, Qureshi AM, Sachdeva R, Maskatia SA, Justino H, Goldberg DJ, Mozumdar N, Whiteside W, Rogers LS, et al. Outcomes after decompression of the right ventricle in infants with pulmonary atresia with intact ventricular septum are associated with degree of tricuspid regurgitation: results from the congenital catheterization research collaborative. Circ Cardiovasc Interv. 2017;10:e004428–004438. doi: 10.1161/CIRCINTERVENTIONS.116.004428

17. Luca AC, Lozneanu L, Miron IC, Trandafir LM, Cojocaru E, Paduret IA, Mihaila D, Leon-Constantin MM, Chiriac S, Iordache AC, et al. Endocardial fibroelastosis and dilated cardiomyopathy - the past and future of the interface between histology and genetics. Rom J Morphol Embryol. 2020;61:999–1005. doi: 10.47162/RJME.61.4.02

18. Crucean A, Alqahtani A, Barron DJ, Brawn WJ, Richardson RV, O’Sullivan J, Anderson RH, Henderson DJ, Chaudhry B. Re-evaluation of hypoplastic left heart syndrome from a developmental and morphological perspective. Orphanet J Rare Dis. 2017;12:138–147. doi: 10.1186/s13023-017-0683-4

19. Pesevski Z, Kvasilova A, Stopkova T, Nanka O, Drobna Krejci E, Buffinton C, Kockova R, Eckhardt A, Sedmera D. Endocardial fibroelastosis is secondary to hemodynamic alterations in the chick embryonic model of hypoplastic left heart syndrome. Dev Dyn. 2018;247:509–520. doi: 10.1002/dvdy.24521

20. Luo G, Liu A, Wang KL, Sun Y, Chen TT, Liu N, Pan SL. [No surgical intervention in neonatal period of pulmonary stenosis or atresia with intact septum after fetal pulmonary valvuloplasty: two cases report]. Zhonghua Er Ke Za Zhi. 2019;57:891–893. doi: 10.3760/cma.j.issn.0578-1310.2019.11.017

21. Xu X, Friehs I, Zhong Hu T, Melnychenko I, Tampe B, Alnour F, Iascone M, Kalluri R, Zeisberg M, Del Nido PJ, et al. Endocardial fibroelastosis is caused by aberrant endothelial to mesenchymal transition. Circ Res. 2015;116:857–866. doi: 10.1161/CIRCRESAHA.116.305629

22. Kuhle H, Cho SKS, Barber N, Goolaub DS, Darby JRT, Morrison JL, Haller C, Sun L, Seed M. Advanced imaging of fetal cardiac function. Front Cardiovasc Med. 2023;10:1206138–1206153. doi: 10.3389/fcvm.2023.1206138

23. Enzensberger C, Graupner O, Fischer S, Meister M, Reitz M, Gotte M, Muller V, Wolter A, Herrmann J, Axt-Fliedner R. Evaluation of right ventricular myocardial deformation properties in fetal hypoplastic left heart by two-dimensional speckle tracking echocardiography. Arch Gynecol Obstet. 2023;307:699–708. doi: 10.1007/s00404-022-06857-x

